# How super-spreader cities, highways, hospital bed availability, and dengue fever influenced the COVID-19 epidemic in Brazil

**DOI:** 10.1101/2020.09.19.20197749

**Authors:** Miguel A.L. Nicolelis, Rafael L. G. Raimundo, Pedro S. Peixoto, Cecília S. de Andreazzi

**Author notes:** Corresponding author: Miguel A. Nicolelis, MD, PhD, Box 103905 Dept. of Neurobiology, Duke University, Durham, NC 27710, Email –.

## Abstract

Although its international airports served as the country’s main entry points for SARS-CoV-2, the factors driving the uneven geographic spread of COVID-19 cases and deaths in Brazil remain largely unknown. Here we show that four major factors likely accounted for the entire dynamics of COVID-19 in Brazil. Mathematical modeling revealed that, initially, the “super-spreading city” of São Paulo accounted for roughly 80% of the case spread in the entire country. During the first 3 months of the epidemic, by adding only 16 other spreading cities, we accounted for 98-99% of the cases reported in Brazil at the time. Moreover, 26 of the major Brazilian federal highways accounted for about 30% of SARS-CoV-2’s case spread. As cases accumulated rapidly in the Brazilian countryside, the distribution of COVID-19 deaths began to correlate with a third parameter: the geographic distribution of the country’s hospital intensive care unit (ICU) beds, which is highly skewed towards state capitals where the epidemic began. That meant that severely ill patients living in the countryside had to be transported to state capitals to access ICU beds where they often died, creating a “boomerang effect” that contributed to the skew of the geographic distribution of COVID-19 deaths. Finally, we discovered that the geographic distribution of dengue fever, amounting to more than 3.5 million cases from January 2019 to July 2020, was highly complementary to that of COVID-19. This was confirmed by the identification of significant negative correlations between COVID-19’s incidence, infection growth rate, and mortality to the percentage of people with antibody (IgM) levels for dengue fever in each of the country’s states. No such correlations were observed when IgM data for chikungunya virus, which is transmitted by the same mosquito vector as dengue, was used. Thus, states in which a large fraction of the population had contracted dengue fever in 2019-2020 reported lower COVID-19 cases and deaths, and took longer to reach exponential community transmission, due to slower SARS-CoV-2 infection growth rates. This inverse correlation between COVID-19 and dengue fever was further observed in a sample of countries around Asia and Latin America, as well as in islands in the Pacific and Indian Oceans. This striking finding raises the intriguing possibility of an immunological cross-reactivity between DENV serotypes and SARS-CoV-2. If proven correct, this hypothesis could mean that dengue infection or immunization with an efficacious and safe dengue vaccine could produce some level of immunological protection for SARS-CoV-2, before a vaccine for SARS-CoV-2 becomes available.

## INTRODUCTION

Barely 6 months after its first report of a COVID-19 case, on February 26, 2020, Brazil recorded the staggering tally of more than 4,000,000 cases and 125,000 deaths [1] as a consequence of the rampant SARS-CoV-2 epidemic that raged through the entire country. Those numbers ensure that, by September 16^th^, Brazil was the third most affected country in the world, right behind the United States and India in terms of both accumulated COVID-19 cases, and second only to the US in terms of deaths [1].

By early March, it became clear that the country’s international airports, located mainly in large state capital cities on the Brazilian Atlantic coast (with only three main exceptions: Brasilia, Belo Horizonte and Manaus) had been the main entry points of SARS-CoV-2 into the country [2]. However, despite the fact that the main genotypes arriving and spreading through the country were rapidly identified [3], the routes taken by SARS-CoV-2 to reach the entire Brazilian territory remained mysterious until now. In addition, the heavily skewed and heterogeneous spatial distribution of COVID-19 cases throughout the country’s five official regions (North (NO), Northeast (NE), Central-West (CO), Southeast (SE) and South (S)), even after six months of an out of control epidemic, as well as the discrepancy between cases and death distributions caught our attention (Figure 1). The focus of this study, therefore, was identifying the key factors that could account for these uneven spatial distributions and, as a second step, explaining how the five Brazilian regions, or even individual states, exhibited quite remarkable differences in key epidemiological indicators (e.g. incidence, rate of growth, date of arrival of cases, and mortality) during the first 6 months of the SARS-CoV-2 Brazilian epidemic.

**Figure 1.**
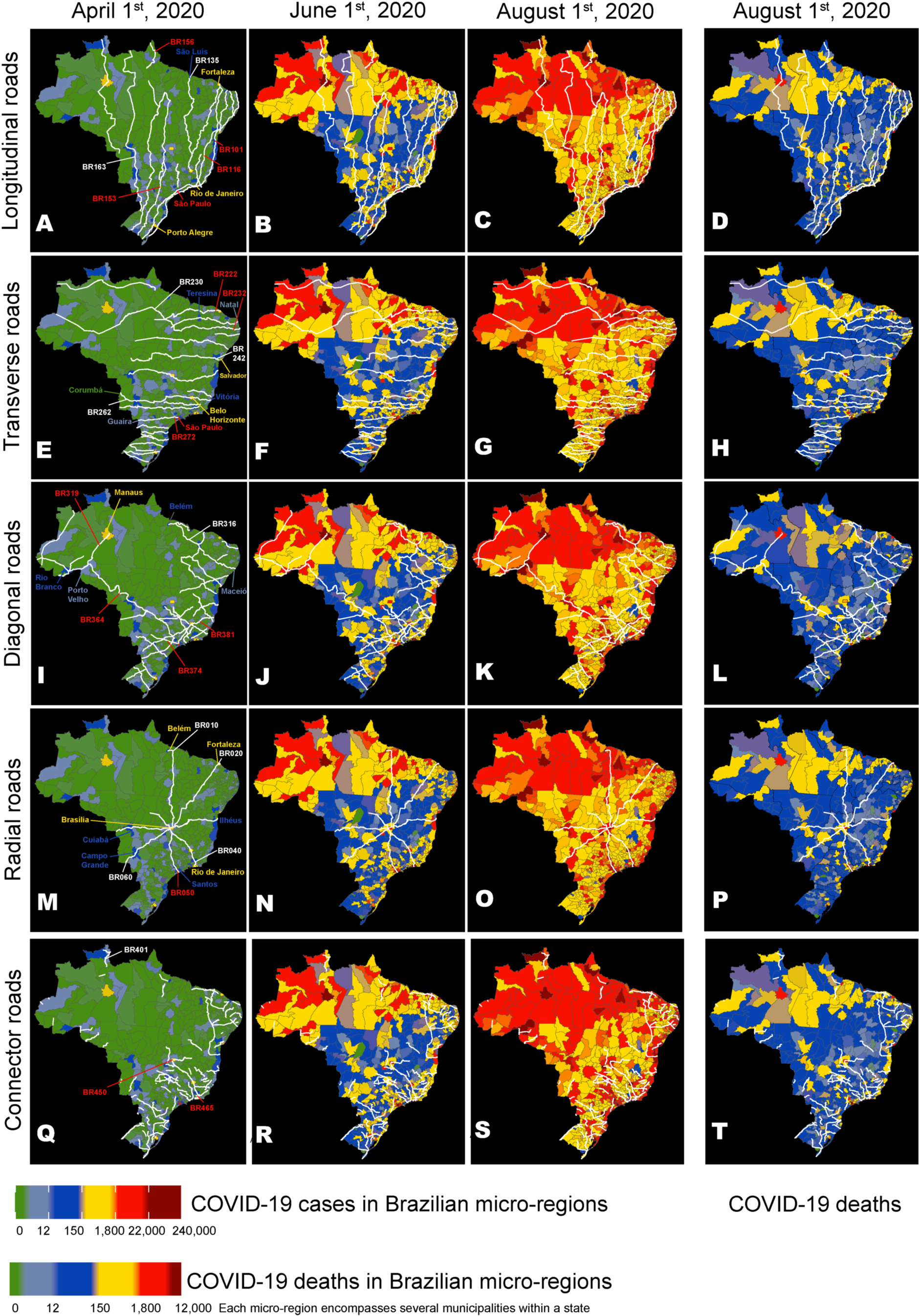
Maps of Brazil were used to represent the routes of the main longitudinal **(A-D)**, transversal **(E- H)**, diagonal **(I-L)**, radial **(M-P)**, and connector **(Q-T)** federal highways, as well as the evolution of the geographic distribution of COVID-19 cases on three dates (April 1^st^, June 1^st^, and August 1^st^), and the distribution of COVID-19 deaths on August 1^st^ (**D**). Overall, a group of 26 highways (see text) from all five road categories contributed to approximately 30% of the COVID-19 case spreading throughout Brazil. The numbers of some of these spreading highways are highlighted in red. Notice how many hotspots (red color) for COVID-19 cases occur in micro-regions containing cities that are located along major highway routes like BRs 101, 116, 222, 232, 236, 272, 364, 374, 381, 010, 050, 060, 450, and 465. Although the distributions for COVID-19 cases and deaths were correlated, geographic discrepancies between the two distributions can be clearly seen by comparing them on August 1^st^ (**C and D**). A color code (See Figure bottom) ranks Brazilian micro-regions (each comprising several tows) according to their number of COVID-19 cases and deaths.

## RESULTS

Figure 1 compares the distributions of all cases and deaths for all Brazilian 5,570 cities from April 1st until August 1st, 2020. Simple visual inspection of these distributions reveals striking spatial patterns in each of them and also a clear dissimilarity. For instance, while by August 1^st^ most of the country was reporting a high number of COVID-19 cases, a larger incidence of fatalities was concentrated on the coastal state capitals and medium-size interior towns (see Figure 1 C and D). To account for such features, we first analyzed the spatial spread of COVID-19 cases and deaths over time through the extensive network of highways that crisscross the whole Brazilian territory, including the vast rain forest of the north. Figure 1A-T illustrates the temporal evolution of the spatial spread of COVID-19 cases by Brazilian micro-regions (each containing several towns) plotted on top of the main routes taken by all longitudinal (north-south, Fig. 1 A-D), transversal (east-west, Fig. 1 E-H), diagonal (Fig. 1 I-L), radial (Fig. 1 M-P), and connector (Fig 1. Q-T) Brazilian federal highways. Beginning with the early phase of the epidemic (April 1st), one can easily spot the spread of COVID-19 cases across the cities either crossed or located near the routes of two major longitudinal highways (BR 101 and BR 116, Fig. 1 A-D) that run from the southern-most state of the country, Rio Grande do Sul (RS), to the north coast states of the NE region. Subsequent snapshots in time (June 1^st^ and August 1^st^) clearly show COVID-19 cases climbing in cities along other major highways, which became hotspots for the epidemic. Following a comprehensive correlational analysis, we observed that a set of 26 federal highways significantly contributed to approximately 30% of the initial COVID-19 spread throughout Brazil (see Supplementary Tables 1). In addition to BRs 101 and 116, these included other longitudinal (BRs 153, 156, Fig 1 A-D), transversal (BRs 222, 226, 232, 272, Fig. 1 E-H), diagonal (BR 316, 319, 324, 364,374, 381, Fig. 1 I-L), radial (10, 20, 40, 50, 60, Fig. 1 M-P), and connector (BR 401, 408,425, 447, 448, 450, 460, Fig 1. Q-T) federal highways. Similarly, a set of federal highways(BRs 101, 116, 222, 232, 272, 308, 319, 374, 381, 20, 40, 50, 408, 447, 450, and 465) was highly correlated with the distribution of COVID-19 deaths across the whole country (Supplementary Table 2).

Next, we focused on identifying the major Brazilian cities contributing to COVID-19 case spread through the Brazilian highway grid. Mathematical modeling (see Methods) revealed that, during the first 3 weeks of the epidemic (from the last week of February to mid-March), by itself the city of São Paulo, which is situated near the largest Brazilian international airport and is responsible for the highest highway traffic flow in the country, was responsible for the spread of more than 80% of the original cases that found their way throughout Brazil (Figure 2A). Because of such a staggering initial contribution, and the fact that it never dropped below 30% for the next 3 months, São Paulo clearly became the main Brazilian super-spreader city of the SARS-CoV-2 epidemic.

**Figure 2.**
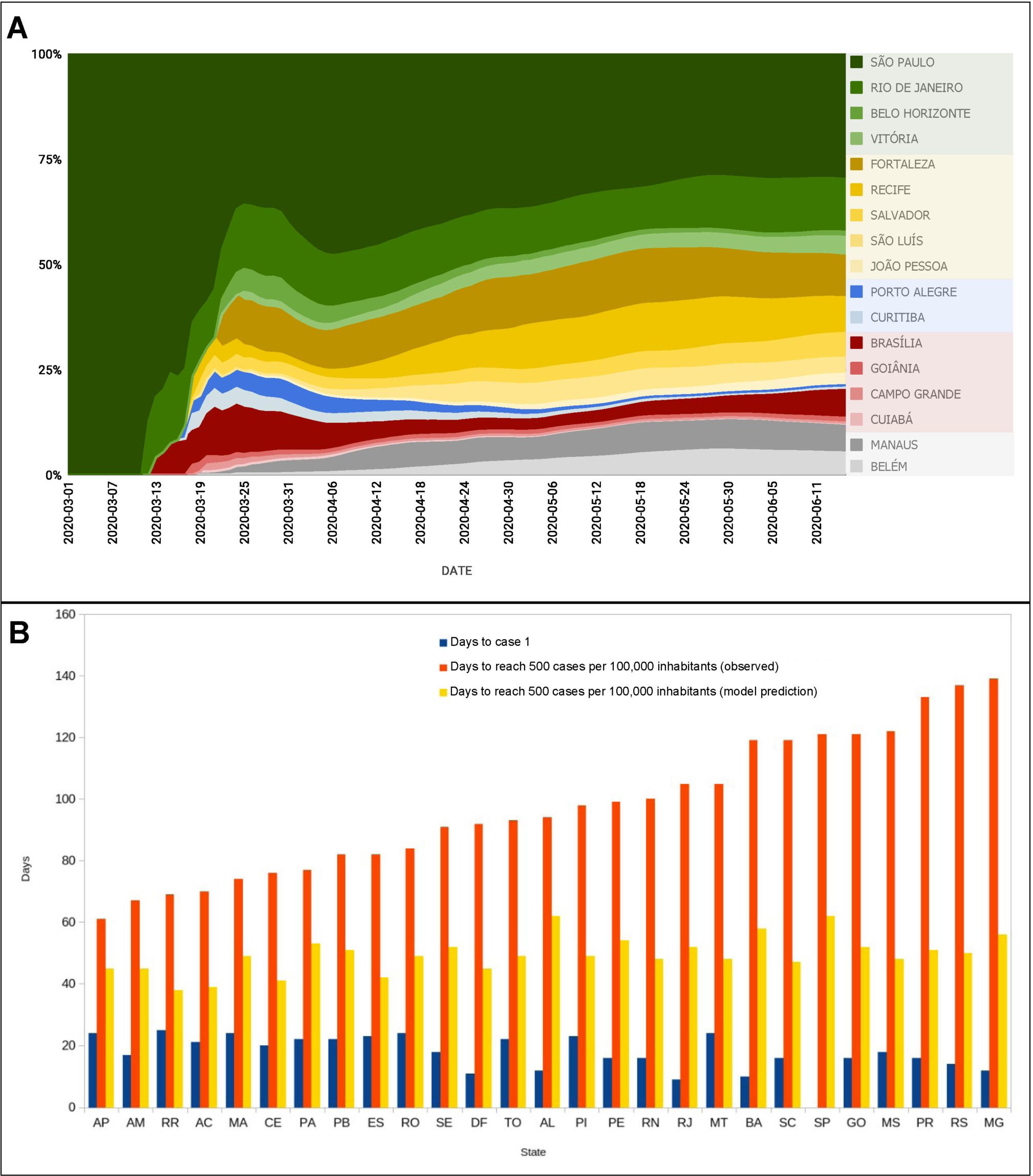
**(A)** Individual contribution of the 17 state capital cities that were responsible for 98% of spreading of COVID-19 cases for the 5570 Brazilian municipalities, from March 1^st^ to June 11^th^. Notice how São Paulo contributed to more than 80% of all case spreading during the first weeks of March. Throughout the period until June 11^th^, São Paulo’s contribution never decreased below 30%. For that reason, the city was labeled as the COVID-19 super-spreader Brazilian city. Notice also the high contribution of Rio de Janeiro, Brasilia, and five state capitals in the Northeast region: Fortaleza, Recife, Salvador, São Luís, and João Pessoa. Manaus and Belém were the largest spreading cities in the North (Amazon) region and Porto Alegre and Curitiba the most important in the South region. During this period, the contributions of Goiânia, Campo Grande and Cuibá, in the Central-West region were the largest in their region but much smaller when compared to other regions and their spreaders. **(B)** Bars represent the day the first COVID-19 case (blue bars) was officially reported in each state (using São Paulo’s first case on February 26^th^, 2020 as the 0 reference), the number of days estimated by a mathematical model for each state to reach 500 cases per 100,000 inhabitants (yellow bars), and the days in which each of Brazilian states actually reach the mark of 500 cases per 100,000 inhabitants (orange bars). Notice how much longer it took for states like MT, BA, SC, SP, GO, MS, PR, MG to reach the 500/100,000 milestone when compared to states like AP, AM, RR, AC, PA, and TO in the North region, MA, CE, PB, PI, SE, AL, and RN in the Northeast region, ES in the Southeast region, and DF in the Center-West region.

Following this initial 3-week epidemic period, other major Brazilian cities began to contribute their share to the spread of COVID-19 cases throughout the country. Thereafter, the cities of Rio de Janeiro (SE region), Belo Horizonte (SE), Fortaleza (NE), Recife (NE), São Luís (NE), João Pessoa (NE), Porto Alegre (S), Curitiba (S), Brasilia (CO), and Manaus (NO) all made significant contributions. Thus, during the first three months of the pandemic, by considering only the top 17 spreading cities, and the highways highlighted above, we were able to account for the spread of about 98-99% of the COVID-19 cases reported in Brazil at the time.

Although the distributions of COVID-19 cases and deaths were significantly correlated (r = -0.886, p< 0.0001), our correlation analysis revealed the existence of an unaccounted residual. This meant that the distribution of deaths (Figure 1D) could not be explained solely by the origin of the cases (i.e. the city in which the person was originally infected). Instead, to account for this residual we had to bring to the foreground what soon became another fundamental factor in the Brazilian COVID-19 epidemic: the geographic distribution of intensive care unit (ICU) beds across the country.

In Brazil, the vast majority of tertiary hospitals, and hence, the largest share of intensive care unit beds is located in state capitals, their metropolitan areas, and a handful of mid-size towns in the interior of each state. By tracking the flow of COVID-19 cases since the beginning of April, and taking into account patient admissions in ICUs nationwide, we were able to identify, in mid-June, a very peculiar flow of people all over Brazil (Figure 3A). As mentioned above, during the initial stages of the epidemic, COVID-19 cases began to grow rapidly in the state capitals where major international airports were located. As cases increased there, a considerable number of infected people began eventually moving towards the vast Brazilian interior through the highway grid. Once they reached their destinations in the countryside, these infected people likely became responsible for community transmission in smaller towns along the roads and immediate vicinities. As community transmission began to happen in earnest, and case numbers rose rapidly in the countryside towns, a growing number of severely ill patients began to overwhelm smaller local hospitals that lacked enough qualified personnel and ICU beds to manage such an unusually high demand for critical care. Under these dire circumstances, a large portion of these patients had to be transported to the large state capitals in search of better specialized care and available ICU beds. In some states, like São Paulo, this patient migration was also directed to mid-size towns with large public university hospitals, such as the cities of Ribeirão Preto and Campinas. As shown in Figures 3A and B, this flow of severely ill patients from the countryside to capitals took place all over Brazil, multiple times during the past 6 months. We named the overall phenomenon that created the flow of infected people from state capitals to the interior, and then brought severely ill patients back to the state capitals and large Brazilian cities, “the boomerang effect”.

**Figure 3.**
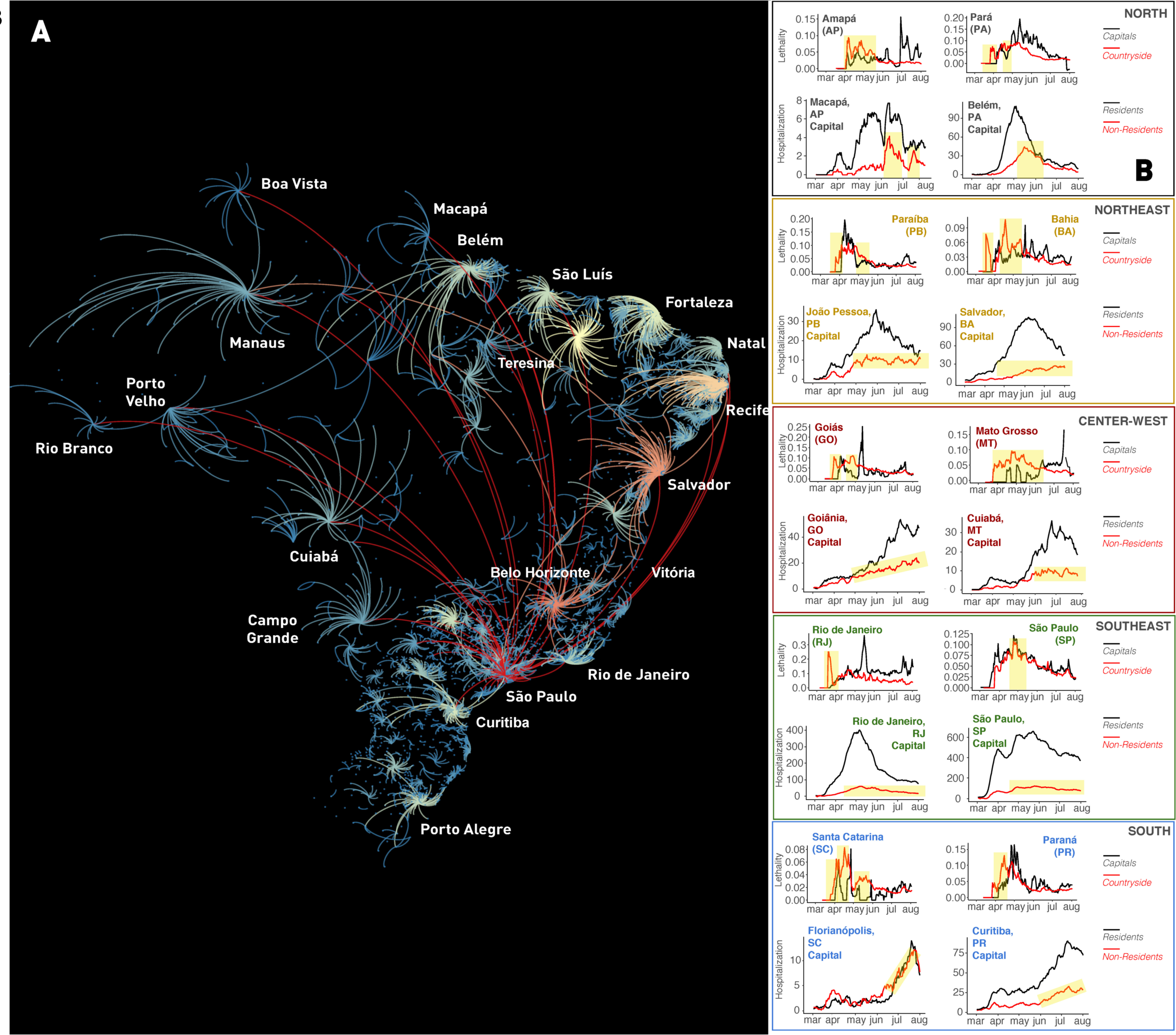
Quantification of the Brazilian “boomerang effect”. **(A)** Representation of all “boomerangs” that occurred around major Brazilian state capitals (see labels for names) and mid-size cities across the whole country. In this map, arcs represent the flow of people from the interior towards the capital. The arc color code represents the number of interior cities that sent severely ill patients to be admitted in hospitals in a capital or mid-size town; red being the highest number of locations, orange and yellow next, while a smaller number of locations are represented in light blue. Most of the flow of people represented in this graph took place through highways. Red arcs likely represent long-distance flow by airplanes. In the Amazon, most of the flow of people towards Manaus occurred by boats through the Amazon river and its tributaries. Notice that again São Paulo appears as the city with the highest boomerang effect, followed by Belo Horizonte, Recife, Salvador, Fortaleza, and Teresina. **(B)** Lethality and hospitalization data, divided for capital and interior (for lethality) and capital resident and non-resident (hospitalization), for a sample of state capitals in all five regions of Brazil. Yellow shading in the lethality graphs represent periods in which more deaths occurred in the interior, in relation to the capital. In the hospitalization graphs, yellow shading depicts periods of increasing admission of people residing in the countryside to the capital hospital system. The overall flow of people from capital to the interior and back to the capital characterized the boomerang effect, targeting the hospital system of the capital city. Notice that boomerang effect was pervasive all over the country, occurring in every Brazilian state.

Figure 3A summarizes all major boomerangs that took place throughout Brazil during the past 6 months. Arcs represent the countryside origins of the largest patient flows towards state capitals and mid-size interior cities for the entire country. Once again, São Paulo emerged as the city with the highest boomerang effect, followed by Belo Horizonte, Recife, Salvador, Fortaleza, and Teresina, all state capital cities (Figure 3A). Boomerangs were so pervasive throughout the country that they triggered major surges in hospital admissions in most state capitals in all Brazilian regions (see yellow highlights in Fig. 3B), leading to peaks of lethality in each of these cities (Figure 3B). Moreover, the boomerang flow was not restricted to roads and highways. For instance, in the Amazon rain forest, severely ill people were transported by boats of all sorts using the large rivers of the North region, from many small riverside communities, towards the two largest Amazon cities, Manaus and Belém (see Figure 3A).

At this point, we decided to test whether the skewed geographic distribution of ICU beds across the country could account for the death distribution residual we described above. Figure 4A illustrates the spatial distribution of ICU beds across all of Brazil. Once this distribution was plotted on top of the COVID-19 death distribution (Figure 4B), the two matched almost perfectly. Thus, when the distribution of ICU beds was summed to the distribution of COVID-19 cases, we obtained a high correlation with the COVID-19 death distribution (R^2^ = 0.846, p = 0.0001). In other words, independently of their original residence, either interior towns or large cities, a significant number of people died in the state capitals and mid-size cities where tertiary hospital facilities and ICU beds were highly concentrated. Therefore, as a result of the boomerang effect, large numbers of severely ill patients had to migrate to larger cities and, eventually, a high fraction of them perished there. Combined with the deaths of the residents of large cities, the widespread boomerang contributed decisively to the geographic skewing of the COVID-19 death distribution in all of Brazil.

**Figure 4.**
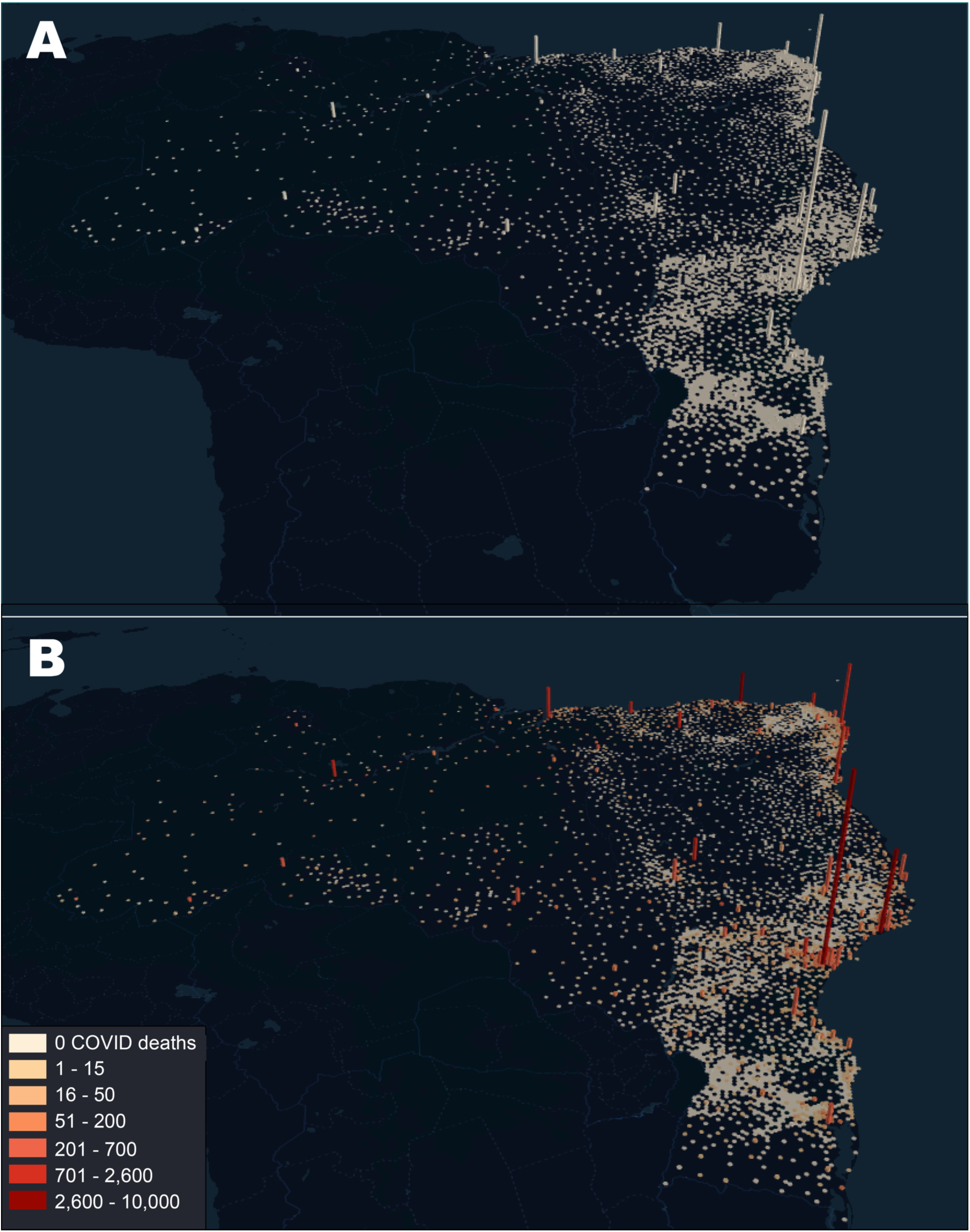
**(A)** Distribution of ICU beds across all Brazil. Bar height is proportional to the number of ICU beds in each city. Notice how the coastal state capitals accumulate most of the ICU beds in the whole country, with much fewer beds available in the interior of most states. The city of São Paulo exhibits the larger number of ICU beds in the whole country. **(B)** Superimposition of the COVID-19 death distribution (color code legend on the left lower corner) on top of the ICU bed distribution as seen in (**A**). For each bar, its height represents the number of ICU beds in a city, while color represents the number of deaths that occurred in that city. Again, the city of São Paulo, which has by far the highest number of ICU beds accumulated the highest number of COVID-19 related fatalities, followed by state capitals like Rio de Janeiro, Fortaleza, Brasilia, Salvador, Manaus, Recife, and Belém.

During our analysis, we noticed a peculiar irregularity in the temporal evolution of the COVID-19 case spread across the Brazilian regions. Contrary to the COVID-19 pattern of spatial spread expected by our mathematical simulations (Figure 2B, yellow bars), we noticed that some states, like Paraná (PR), Santa Catarina (SC), Rio Grande do Sul (RS), Mato Grosso do Sul (MS), Mato Grosso (MT), Goiás (GO) Minas Gerais (MG) and Bahia (BA), which reported their initial cases (Figure 2B, blue bars) during the month of March, tended to display a very slow growth of COVID-19 cases thereafter (Figure 2B, red bars). That happened despite the fact that these states were all crossed by major highways and received a great deal of traffic flow from the super-spreading city of São Paulo. Such differential spread in cases further defined the peculiar shape of the distribution of COVID-19 cases across Brazil by June 30^th^ (Figure 5A).

**Figure 5.**
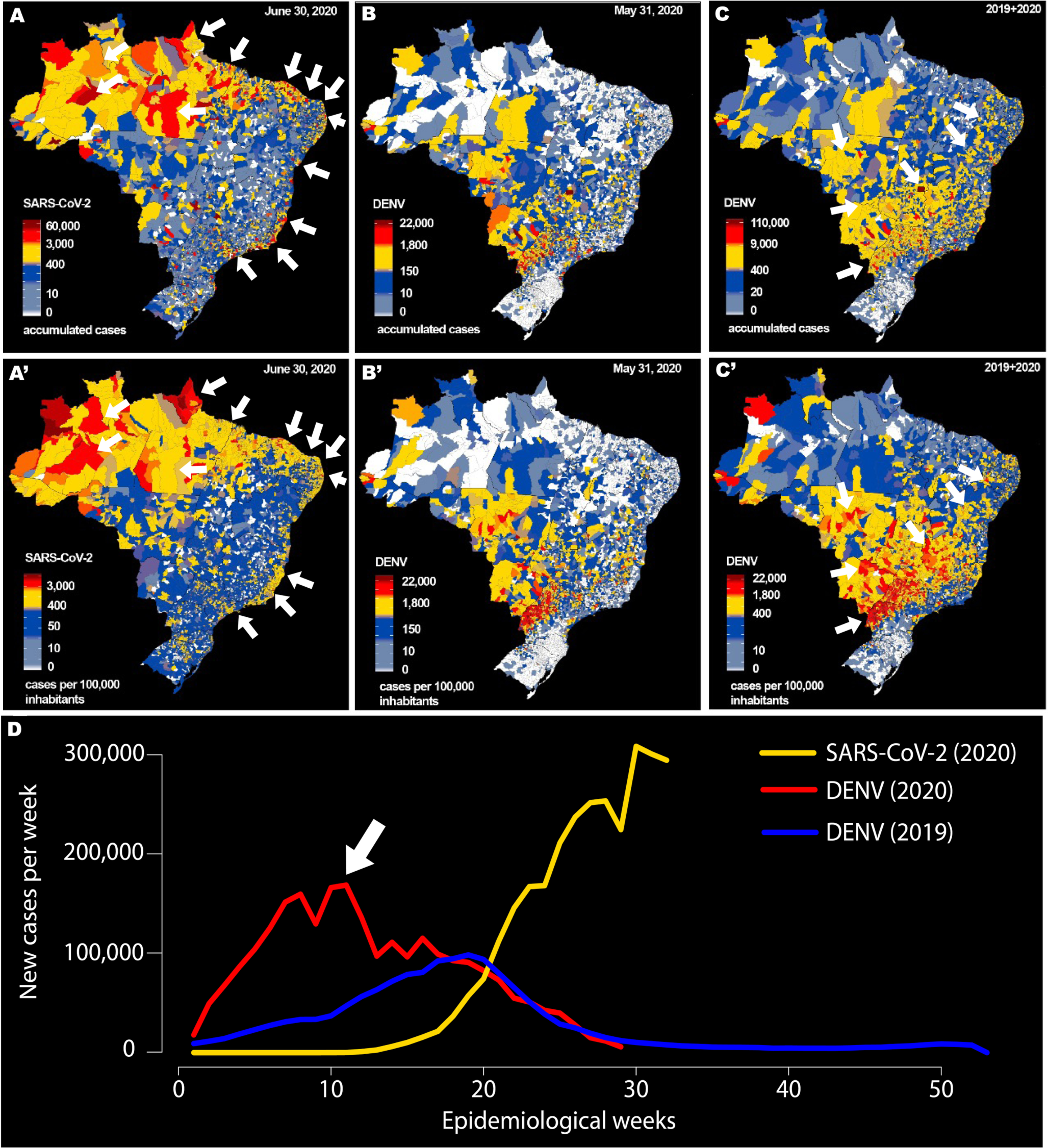
Comparison between the geographic distribution of COVID-19 cases **(A)**, and incidence (**A”**) until June 30^th^, 2020, and dengue fever cases and incidence in Brazil (**B and B’**) until May 31^st^, 2020, and for dengue fever cases and incidence (**C and C’**) summing all data from 2019 and until May 31^st^, 2020. Notice that the COVID-19 and the dengue maps for 2020 and 2019 plus 2020 are rather complementary, with regions in which COVID-19 cases and incidences were very high by late June 30^th^, like the entire North region (see white arrows) and the Brazilian coast (see white arrows), from São Paulo, Rio de Janeiro, and Vitória (SE region), passing through all major capitals of the Northeast region (Salvador, Aracaju, Maceió, Recife, João Pessoa, Natal, Fortaleza and São Luís, all the way to Belém in the North region. On the other hand, the highest number of dengue cases and incidence (C) are distributed over the west region of the Paraná, São Paulo, and Mato Grosso and Mato Grosso do Sul, Goiás, and Brasilia, the country’s capital (see white arrows in C and C’). The states of Bahia (NE region) and Minas Gerais (SE region) also showed very high dengue incidence levels (see white arrows). Comparison of the evolution of COVID-19 (yellow line), dengue 2019 and dengue 2020 new cases per epidemiological week. Notice how dengue fever cases in 2020 began to drop quickly when COVID-19 cases start to grow rapidly in Brazil (see white arrow). That happened even though dengue fever cases in 2020 grew at a much higher rate than in 2019, leaving the Brazilian Ministry of Health to predict that in 2020 the dengue epidemic would be much worse.

By the end of June (Figure 5A and A’), outside the vast area affected in the NO region, a series of vast “empty COVID-19” regions were identified in the central-west (CO) and southeast (SE) regions and all over most of the transitional inland territory, immediately westward from the Atlantic coastal cities (see white arrows in Figure 5A and A’), which by then had all become major COVID-19 hotspots. This coastal hotspot line extended all the way from São Paulo (SE) to São Luís (NE), passing through the metropolitan area of large capital cities like Rio de Janeiro, Salvador, Aracaju, Maceió, Recife, Joao Pessoa, Natal, and Fortaleza (see white arrows in Figure 5A and A’).

After ruling out a series of other factors, we decided to search the Brazilian Ministry of Health databases and its regular epidemiological bulletins for any potential interfering factors that could account for this peculiar skewed shape. Figure 5B-C and B’-C’ reveals the rather surprising finding of our search. In Figure 5A and A’ we reproduce the Brazilian map with the distribution of COVID-19 cases (Figure 5A) and incidence (Figure 5A’) by municipality on June 31st. Next to it, we display the Brazilian maps depicting the spatial distributions of confirmed dengue fever cases (1,337,095 in 2020) and incidence in 2020 (Figure 5B and B’), and the total sum of dengue cases (3,585,665 cases) and incidence for 2019-2020 (Figure 5C and C’) for all Brazilian towns.

Visual inspection of these maps clearly indicated that they were complementary; i.e. areas in which there was a scarcity of COVID-19 cases were equivalent to regions in which a large concentration of dengue fever cases had occurred during the Brazilian dengue epidemic of 2019-2020 (see white arrows in Figure 5C and C’). Indeed, all the abnormalities observed in the COVID-19 case rate of growth and the delays in the COVID-19 case curves of states like PR, SC, RS, GO, MS, MT, MG and BA (Figure 2B) could now be explained by the geographic distribution of large concentrations of dengue fever cases in these states.

Figure 5D plots the evolution of dengue fever cases in Brazil in 2019 and 2020, together with the equivalent curve for COVID-19 cases in 2020. Notice that a precipitous fall in the number of dengue fever cases began by epidemiological week (EW) 11 (03/08– 03/14/2020, see white arrow). This was paralleled by a concurrent growth of COVID-19 cases in Brazil. In fact, the dengue fever cases in 2020 dropped much earlier than expected according to its seasonal pattern [4], as illustrated by the curve of cases obtained in 2019.

To further characterize the potential interaction between dengue fever and COVID-19, we plotted the incidence of COVID-19 cases and deaths, as well as the rate of growth of COVID-19 cases and the number of days required to reach 1,000 COVID-19 cases per 100,000 inhabitants per state, as a function of the percentage of the population in each state showing positive IgM antibodies for dengue fever, as reported by the Brazilian Ministry of Health (Figure 6A-D). Further analysis revealed the existence of a highly significant inverse exponential correlation between COVID-19 cases (r = -0.659, p< 0.0001) and deaths (r= -0.514 p< 0.006), as well as COVID-19’s infection growth rate (r= -0.662, p< 0.0001) and the percentage of people exhibiting positive IgM to the dengue virus (DENV) in each state. These inverse correlations grew over time until reaching a peak around July 1^st^, 2020. Furthermore, when the number of days needed for reaching 1000 cases/100,000 inhabitants were plotted against the DENV IgM levels per state, we observed a significant linear positive correlation (r= 0.594, p< 0.001). Essentially this analysis revealed that states that had higher incidence of dengue fever during the 2020 dengue epidemic were less likely to exhibit COVID-19 cases and deaths, showed slower rates of growth of COVID-19 infections, and hence took longer to accumulate COVID-19 cases. In Figure 6A-F we also highlighted the dengue serotypes (DENV 1-4) that were identified in each Brazilian State (see Brazilian Map on the insert of Figure 6A). Although we did not explore this variable further, it is interesting to notice that the combination of DENV 1-2 was the most prevalent in most of the country in 2020.

**Figure 6.**
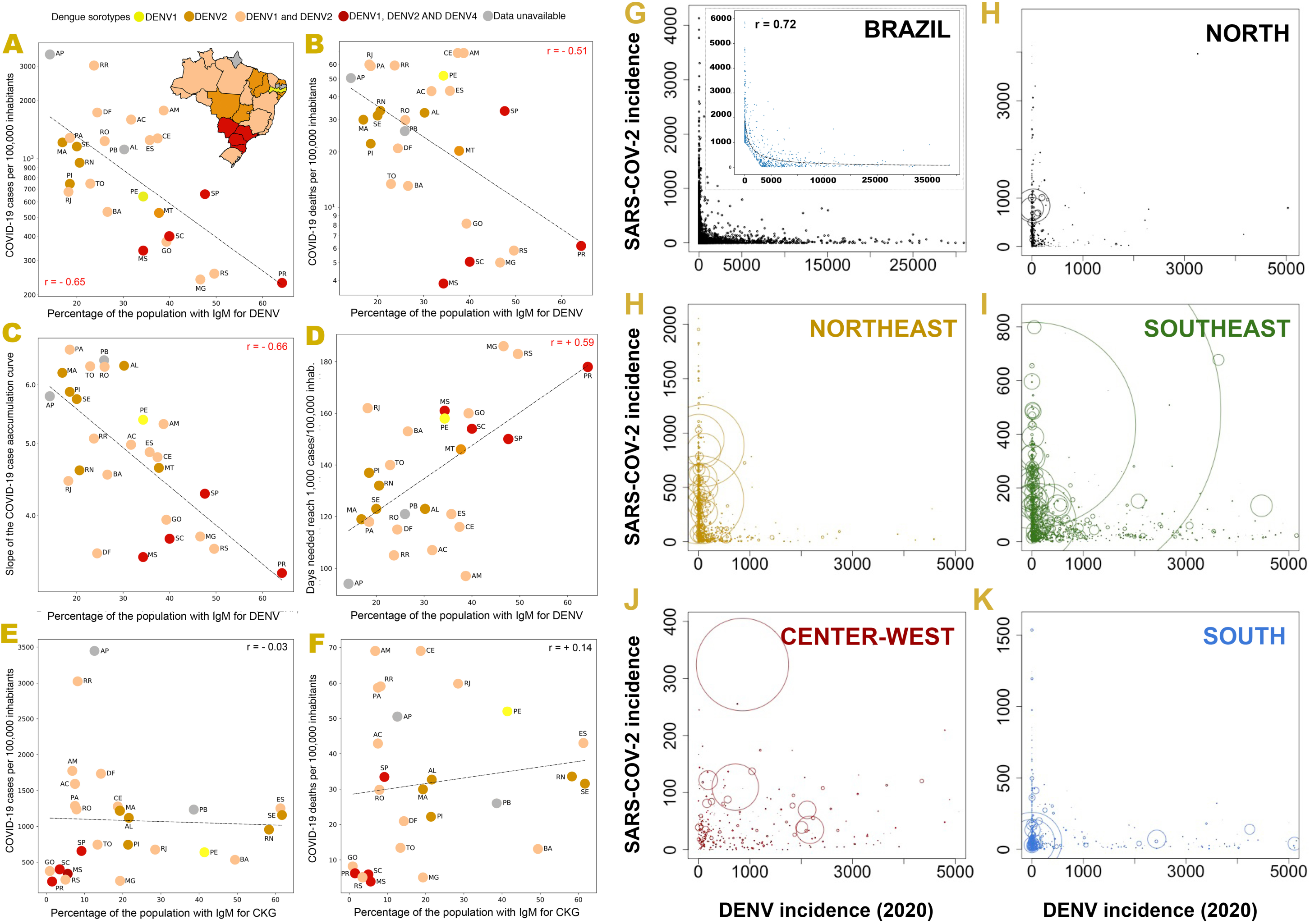
(**A-F**) Inverse exponential correlations between COVID-19 case incidence per 100,000 per inhabitants (**A**, r = -0.659, p<0.0001), COVID-19 death incidence (**B**, r = 0.51, p <0.006), rate of growth of COVID-19 cases (**C**, r = -0.66, p < 0.0001) against the percentage of state population with positive IGM for dengue fever. (**D**) Positive correlation between number of days to reach 1,000 COVID-19 cases per 100,000 inhabitants and percentage of state population with positive IGM for dengue fever. (**E and F**) Lack of correlation between COVID-19 case. Notice that the Y axes in the plots **A-C** are in logarithmic scale. There was no correlation between COVID-19 case (**E**) (r = -0.03, p = 0.84) and death incidence (**F**) (r = 0.141, p = 0.49) as a function of the percentage of state population with IgM for chikungunya virus. (**G-L**) Scatter plots depict the inverse interaction between COVID-19 incidence and the 2020 dengue fever incidence for all 5,570 Brazilian municipalities (**G**) and divided per the North (**H**), Northeast (**I**), Southeast (**J**), Center-West (**K**), and South (**L**) regions. In each plot, the diameter of circles represents city population. These scatter plots reveal that cities with high COVID-19 incidence exhibited very low dengue fever incidence and vice-versa. This inverse relationship is better described in a subset of 799 cities from all regions using a hyperbolic fit (y = -155.9+1349476.6/(x+1000)) (r = 0.72, p = 0) (**G’**). For this analysis, we disregarded cities with small incidence in both DENV (x) and COVID (y), following the threshold of y + [max(y)/max(x)] x < 1,000 cases per 100,000 inhabitants. That explains the triangular edge at the bottom of the graph. A similar hyperbolic fitting was also obtained for 1,466 cities when the incidence cutoff was reduced to y + [max(y)/max(x)] < 500 cases per 100,000 inhabitants (r = 0.55, p = 0.0).

As a control measure, we repeated all the above analysis (see Figure 6E and F) using serological data obtained during the same 2020 period for patients diagnosed with chikungunya, a virus mainly transmitted by the same insect vectors of dengue fever (*Aedes aegypti and Aedes albopictus*), and which is also endemic in Brazil, albeit at much lower levels. No significant correlation to any COVID-19 epidemiological parameter was found with the percentages of people displaying positive IgM for chikungunya (CKG) in all Brazilian states (Figure 6E, COVID-19 case incidence and CKG IgM, r= -0.03, p = 0.84; COVID-19 death incidence and CKG IgM, r= 0.141, p = 0.49).

To further investigate the potential relationship between dengue fever and COVID-19, we plotted the incidence of COVID-19 versus dengue in 2020, for all 5,570 Brazilian cities; all together (Figure 6G) and according to the specific region to which they belonged, such as the North (Fig. 6H), Northeast (Fig. 6I), Southeast (Fig. 6J), Center-West, (Fig. 6K), and South region (Fig. 6L)). These latter scatter plots revealed a clear inverse pattern of interaction for each region with clear interregional differences. For instance, while the NO states had a larger proportion of cities with very high COVID-19 incidence, and very low dengue incidence, the CO states had a distribution that was the mirror opposite: a much larger number of cities with high dengue incidence and lower COVID-19 cases.

While dengue cases were more numerous in the NE, compared to the NO region, this region was dominated by cities with high COVID-19 and low dengue incidences. A larger number of cities with high incidence of dengue, and very low COVID-19 incidence, occurred in the SE and SO regions. By adding to these plots circles to represent the population size of every town per region, we also observed another interesting phenomenon: COVID-19 tended to dominate in larger cities where the dengue incidence was much lower than in mid- or small-size towns, where dengue incidence was much higher and the incidence of COVID-19 much lower.

To further quantify the inverse relationship between COVID-19 and dengue fever incidence, we selected a subset of 799 cities from all regions and tried to describe their relationship using a hyperbolic function (see Methods for details, Figure 6G’). The fitting of a hyperbolic function was very significant (r = 0.72, p = 0.0001). A similar hyperbolic fitting was also obtained for a sample of 1,466 cities (r = 0.55, p = 0.0001).

To test whether the inverse correlation between COVID-19 and dengue key epidemiological indicators observed in Brazil was specific to the country or more general, we collected incidence data for COVID-19 and dengue fever for a sample of 15 countries around the world, notably in Southeast Asia, Latin America, and several islands in the Pacific and Indian Ocean were dengue is known to be very prevalent (Figure 7). When the incidence of COVID-19 versus the incidence of dengue fever for 2019-20 was plotted for these countries, we again obtained a highly significant inverse exponential correlation (r = 0.7794 and p< 0.0006). In other words, the more dengue fever cases a country had during the worldwide dengue epidemic of 2019 and the first few months of 2020, the less COVID-19 cases the country exhibited until July 2020. Basically, this was very similar to the results obtained using the data for the Brazilian states.

**Figure 7.**
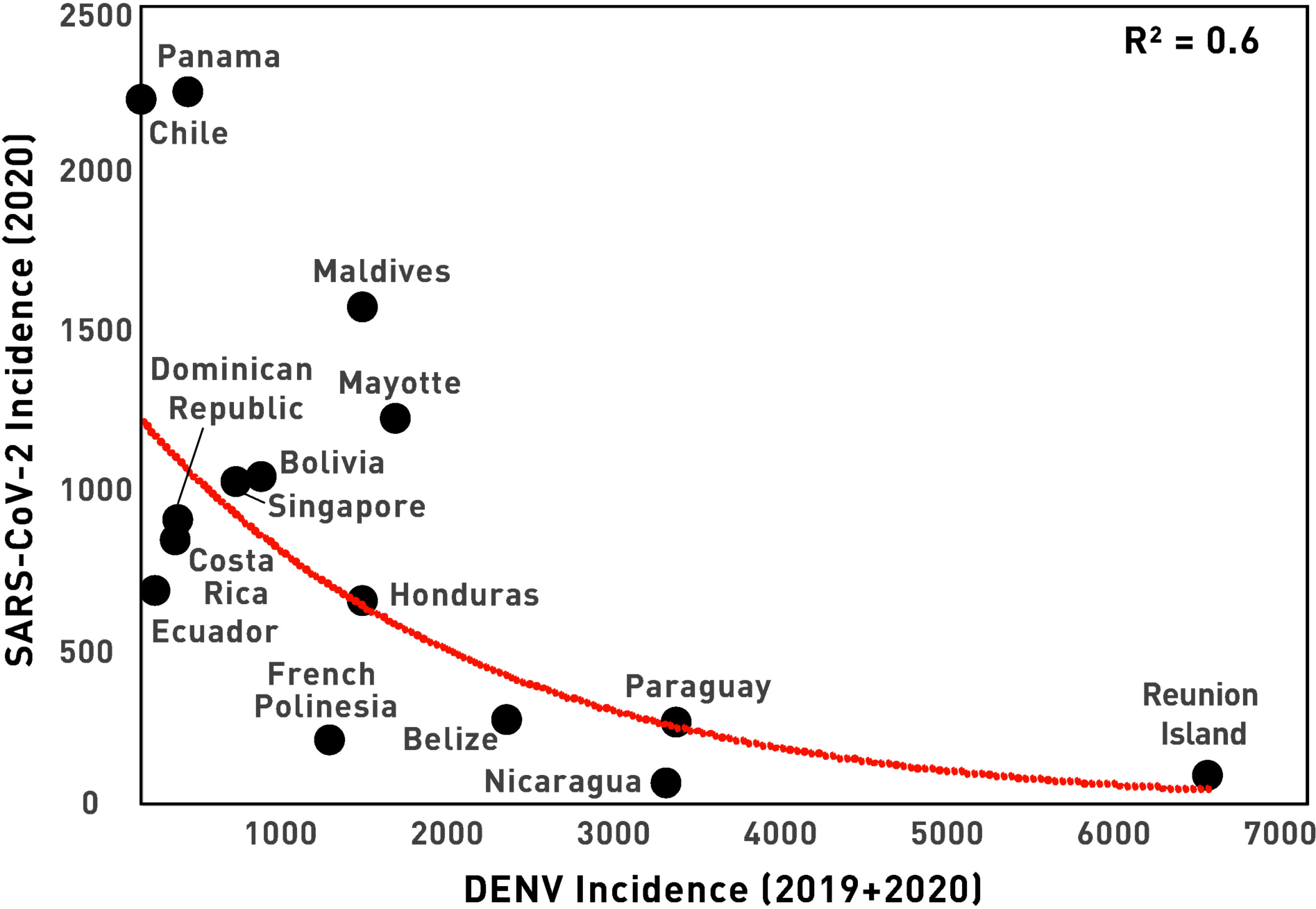
Inverse exponential correlation (R2 = 0.605, r = 0.7794 and p< 0.0006) between COVID-19 case incidence as a function of the dengue fever incidence for a sample of countries in Latin America, Asia, and a few islands in the Pacific and Indian Oceans.

## Discussion

Overall, we identified four major factors that concurrently accounted for most of the dynamics of the COVID-19 pandemic in Brazil. From its original entry at all major Brazilian international airports during the month of March [2], SARS-CoV-2 spread first to the large metropolitan areas of state capitals located next to these airports. From that point on, after community transmission was established and began to rise exponentially in these cities, and given that no major road blocks were implemented during the early months of the epidemic, a small group of these large cities began spreading SARS-CoV-2 to the entire country, through the extensive highway grid that covers all of Brazil. By itself, São Paulo, the city with highest population in Brazil, emerged as the country’s super-spreader city par excellence, accounting for the largest case spreading influence throughout the next 3 months. A small set of other 16 spreading cities contributed to the seeding of initial cases throughout the whole country via a subset of 26 major federal highways. This highway-driven spread was the main mechanism through which initial cases arrived in all Brazilian cities. Thus, in about 30 days SARS-CoV-2 was transported to all five regions of the country, across the north-south axis, a distance of roughly 5,313 kilometers.

Our analysis confirmed yet again the extreme relevance of human mobility in spreading infectious diseases [5-8]. Our data also corroborated, at a national level, a recent analysis of the spread of COVID-19 cases to the interior of the state of Pernambuco, which implicated a major transverse federal highway, BR 232, as well as other smaller state roads [9]. Since Brazil’s air space remained open for international (and national) travel until the end of March, and no travel restrictions were imposed on the main roads leaving the super-spreading city (São Paulo) and other major Brazilian state capitals, Brazilian highways provided transportation for infected people to all parts of the country for a full month after the first case was reported in São Paulo. Thus, by the time (mid- to late-March) state governments began issuing decrees imposing social isolation measures for all people (except for those deemed essential workers), all the pre-conditions for COVID-19 community transmission around the entire country, were already in place. Our analysis revealed that traffic through federal highways alone contributed to 30% of this COVID-19 case spread, but since we did not analyze state and municipal roads like other studies [9], the contribution of roads to the movement and spread of infected people all over Brazil is likely to be much higher.

We also observed that the distribution of COVID-19 related deaths overlapped quite well with the equivalent spatial distribution of ICU beds throughout Brazil. The higher the number of ICU beds in a city, the higher the number of deaths it recorded from March to September 2020. Nationwide analysis of the cause of this overlap, which we named the boomerang effect, revealed that while state capitals, mostly located on the country’s Atlantic coast, provided the main sources of infections to mid- and small-size towns located in Brazil’s vast interior, later on these same interior towns countered this flow by sending hordes of seriously ill patients back to the capitals in search of better hospital infrastructure and available ICU beds. As a result of this gigantic human flow, people from interior towns began to account for a large percentage of patient admissions in both public and private hospitals in state capitals. Thereafter, several of these hospitals in both mid-size towns and state capitals became overwhelmed and some even collapsed altogether, like in the city of Manaus [10] where the boomerang effect was mainly operating through the Amazon River. During a few weeks, ICU bed occupancy reached more than 90% in multiple Brazilian state capitals, an event never before seen in Brazilian medical history [11]. Although most state governments tried to mitigate this crisis by quickly adding new infirmary and ICU beds to their hospitals, the lack of specialized personnel, individual protection and sophisticated medical equipment, such as modern artificial ventilators, reduced the efficiency of such countermeasures. As a result, each ICU bed available in the country accounted for 1.23 deaths, according to our partial correlation analysis.

The Brazilian federal health care system, known as the “Sistema Único de Saúde” (SUS; in English: Unified Health System) was created 32 years ago [12] with the mission to provide free health care to every Brazilian citizen anywhere in the country. Today, SUS constitutes the only option through which 7 out of 10 Brazilians have access to high-quality medical care for free [13]. Yet, the COVID-19 epidemic crisis clearly exposed the inadequacy of the policy of concentrating the largest share of tertiary hospital facilities and ICU beds in a handful of mid-size towns and state capitals throughout Brazil. Although our study did not address this issue directly, its findings suggest that, had the geographic distribution of ICU beds been less skewed toward big cities, many more lives could have been saved throughout the country. Indeed, critically ill patients in less populated areas would have had regional access to ICU beds and therefore would have received quicker treatment and had a better chance for improved clinical outcomes. Regional access would also have reduced the demand on critical resources and specialized medical personnel necessary for transporting such seriously ill patients over long distances to metropolitan hospitals, eliminating the widespread boomerang effect documented here.

By far the most unexpected finding of our study was the discovery that the spatial distribution of dengue fever cases in Brazil, recorded during the period of January 2019 to July 2020, was almost perfectly complementary to the distribution of COVID-19 cases for most of the first 6-month period of the SARS-CoV-2 epidemic. Further analysis revealed that the incidence of COVID-19 cases and deaths, as well as the growth rate of the infection, exhibited a significant inverse correlation with the percentage of state residents exhibiting high IgM levels for DENV in 2020. That meant that in states in which dengue fever had been rampant during the 2019-2020 dengue epidemic, fewer COVID-19 cases were reported, fewer people died of COVID-19, the rate of growth for COVID-19 infection was slower, and more time was needed to reach progressive case incidence thresholds (e.g. 1,000 COVID-19 cases per 100,000 inhabitants) in 2020. Since, we did not find any correlation between the percentage of people in each state exhibiting IgM to CKG virus to any COVID-19 epidemiological indicator, our analysis confirmed the specificity of our findings to the DENV serotypes responsible for dengue fever.

To the best of our knowledge, such inverse interactions between dengue fever and COVID-19 have not yet been described in the literature. Given that the four Flavivirus serotypes that cause dengue fever are not closely related to SARS-CoV-2 whatsoever, this finding was really very striking. Nonetheless, these inverse correlations clearly helped us understand the peculiar shape of the geographic spread of COVID-19 cases, in the early months of the Brazilian epidemic. Basically, the states belonging to the CO and S regions (e.g. PR, MS, MT, GO, RS, SC states), as well as in specific subregions of the states of Minas Gerais and Bahia that had a high incidence of dengue fever cases during 2019-2020, took longer times to reach significant numbers of COVID-19 cases than predicted by our mathematical modelling. Given that the higher the percentage of a state’s population exhibiting IgM to DENV, the longer it took for that state to cross 1,000 COVID-19 cases per 100,000 inhabitants, this finding seems to indicate that the huge 2019-2020 epidemic that produced more than 3.5 million reported cases of dengue fever in Brazil could have contributed significantly to the slowing down of the spatiotemporal spread of COVID-19 in 2020, reducing considerably both the total number of COVID-19 cases and deaths. Since close to 100-400 million people around the world become infected with dengue fever each year [14], this finding could have a significant impact on the management of the current SARS-CoV-2 pandemic.

Assuming that other independent factors that could have contributed independently to the occurrence of such inverse correlations can be ruled out, the most parsimonious interpretation of our findings is that through some, as of yet unknown, mechanism, SARS-Cov-2 and the DENV competed for the same pool of susceptible people and that those who contracted dengue during 2019-2020 may have been protected, to some degree, from infection by SARS-CoV-2. A few further findings support this hypothesis. First, while dengue cases in 2019 had reached a very high number (2,248,570), the Brazilian Ministry of Health alerted to the growing incidence of dengue cases during the first two months of 2020 (in: Boletim Epidemiológico 10 [15]). Yet, starting on EW 11, dengue cases began dropping precipitously in Brazil. That was precisely the time that the COVID-19 epidemic began increasing in most states of the SE, in the entire NO region, and in 8 out of 9 NE states. Research of international reports on dengue fever revealed that a similar dramatic fall in dengue fever cases has happened all over the world around the time the COVID-19 pandemic began, particularly in countries in Southeast Asia and most Latin American nations that experienced a rampant dengue fever epidemics in 2019 [16]. Thus, whenever COVID-19 arrived in a given country in early 2020, the occurrence of new dengue fever cases tended to diminish quickly and then almost disappear from the record. This worldwide phenomenon further strengthened our finding of a potential inverse relationship between key epidemiological parameters describing the concurrent COVID-19 and dengue fever epidemics taking place in Brazil in 2019-2020. Seen in this context, there is a possibility that widely popularized success stories of COVID-19 management, like those from Vietnam and other Southeast Asia countries, and even the whole continent of Africa [17], may owe, in reality, a great deal of their good fortunes in handling the COVID-19 epidemic to their high dengue fever prevalence in 2019-2020.

In its official epidemiological bulletins issued regularly, the Brazilian Ministry of Health attributed the sudden decline in dengue cases to possible under-notification problems caused by the COVID-19 epidemic [18]. We, instead, propose that at least part of such precipitous drops in both dengue fever cases likely may reflect the fact that SARS-CoV-2 was rapidly outcompeting the dengue Flavivirus serotypes for infecting the same pool of susceptible people across Brazil. Essentially, if a subject became infected with SARS-CoV-2, he/she would not be infected by dengue viruses. Clearly, further epidemiological and immunological studies will be needed to test this hypothesis.

According to our hypothesis, SARS-CoV-2 may have been able to outcompete DENV, and trigger a sudden decrease in dengue fever incidence all over the world after March 2020, primarily because it relies on human-to-human transmission, while dengue viruses depend mainly on mosquitos of the Aedes genus (*Aedes aegypti and Aedes albopictus*) for transmission. Given that the dengue’s vector can only survive under certain climate conditions, like altitudes below 2,000 meters [19-21], SARS-CoV-2 would have a clear competitive advantage when introduced in a common ecological niche, being able to spread faster and infect a larger population of susceptible people, over a much larger territory, particularly in big cities. This seems to be confirmed by the fact that larger Brazilian cities exhibited much higher incidence of COVID-19 than dengue. Conversely, in medium and small cities the dengue virus was able to infect a great fraction of the susceptible population and, according to our hypothesis, may have protected them somewhat from acquiring COVID-19.

This previously unknown “dengue effect” may explain, at least in part, why most mathematical modeling carried out in the early stages of the COVID-19 Brazilian epidemic were significantly off in their predictions of cases and deaths for the country [22, 23]. The same happened with predictions for countries in Africa and Southeast Asia. In the context of our findings, the explanation for this mismatch could be that previously acquired immunity for dengue fever may have protected people from contracting SARS-Cov-2. This was confirmed by the discovery of a highly significant, exponential inverse correlation between the incidence of dengue and COVID-19 when data from several countries in Latin America, Southeast Asia, and along the Pacific and Indian Oceans were pooled and analyzed together. Detailed analysis of these worldwide data will be covered in two other upcoming studies from our group.

In patients infected by one of the serotypes of DENV, IgM titers begin to increase in the bloodstream after the first couple of days of acute illness [24]. They peak around 7-14 days and disappear after 50-60 days [24]. In parallel, the patients’ lymphocyte B cells begin producing IgG antibodies, which also peak around 2 weeks after the onset of symptoms but, different from their IgM counterparts, remain in high titers for much longer periods of time [24]. That explains why patients infected by one of the four DENV serotypes retain long-term immunity to that serotype, albeit not to the other three [25, 26]. Since the IgM lifetime is so short, it seems logical to conclude, therefore, that the highly significant negative correlations we observed in Brazil, between state populations levels of dengue-induced IgM and several epidemiological indicators of COVID-19 infection, was at least concurrently paralleled by the production of higher titers of IgG antibodies in the dengue affected population. This seems to be confirmed by the fact that when we analyzed data from several countries around the world, high inverse correlations were found initially by using only 2019 dengue fever incidence data against 2020 COVID-19 incidence in each country. That further suggests that a single episode of dengue fever could suffice to generate some level of long-term, IgG-mediated cross-immunity to COVID-19.

Heterologous immunity is a well-known phenomenon, particularly between closely related species of parasites, protozoa, bacteria and viruses [27]. However, it has also been documented between unrelated species. Indeed, this phenomenon led to the hypothesis that routine use of the bacillus Calmette-Guérin (BCG) vaccine in certain countries could explain their lower incidence to a variety of viruses [28, 29], and that the BCG-induced trained immunity could even offer a protection against COVID-19 [30]. Cross-reactive specific antibodies, but also T cells, can underlie heterologous immunity [27]. For instance, pre-existing cross-reactivity, mediated by CD4^+^ T cells to SARS-CoV-2 has been reported in 25-50% of people not exposed to the new coronavirus [31, 32]. Recently, Mateus et al. reported that a range of such CD4^+^ T cells can exhibit cross-reactivity between SARS-CoV-2 and a series of other coronaviruses that cause the common cold [33].

This year a few reports have indicated the occurrence of serological cross-reactivity between dengue fever and COVID-19 [34-37]. For instance, Lustig et al. reported that in a group of 55 patients who tested positive for SARS-Cov-2, 12 patients tested positive for dengue (nine cases positive IgM, 2 positive IgG, and 1 for both). Moreover, they also reported that 21 (or 22%) out of 95 serum samples of patients diagnosed with dengue fever prior to September 2019 (before the outbreak of the COVID-19) exhibited positive/equivocal SARS-CoV-2 serology targeting the S protein (sixteen IgA and five IgG) [35]. The same authors indicated that in-silico analysis revealed potential similarities between SARS-CoV-2 epitopes in HR2-domain of the spike protein and the envelope-protein of both Zika and dengue viruses [35]. In another report, two patients in Singapore who were diagnosed originally through a serological test as having been infected with a dengue virus, later on proved to have contracted SARS-CoV-2 [37]. A recent study conducted in Brazil showed that, in a pool of 44 patients who had dengue, one case also exhibited a false positive result for two COVID-19 rapid tests [38]. The same study showed that, in another pool of 32 patients who had tested positive for SARS-Cov-2, no one exhibited positive IgG/IgM results for dengue virus [38]. Finally, Nath et al. observed that out of 13 serum samples positive for dengue antibody collected in 2017 (before the COVID-19 pandemic), five produced false positive for SARS-CoV-2 when rapid IgG/IGM tests were employed [36]. While the authors of these studies interpreted their results as indicating specificity problems with COVID-19 serology tests, in light of our results, we believe that these findings may provide the first immunological-based evidence of a potential genuine cross-reactivity between DENV and SARS-CoV-2. Moreover, since no study so far has focused on the potential role of T cells in mediating cross-reactivity between the DENV and SARS-CoV-2 viruses, the level of heterologous immunity between these two viral families may be higher than shown by antibody-based studies. Thus, based on these preliminary reports and our own epidemiological findings, we postulate that cross-reactivity, both cellular and humoral, may occur between one or more DENV serotypes that cause dengue fever and COVID-19.

But what is the best antigen candidate that could produce such an immunological cross-reactivity between the dengue virus and SARS-CoV-2? Two recent studies suggest that nucleocapsid proteins in both virus families could fulfill this role. In a drug testing study, Mukherjee and Roy claimed that two anti-viral drugs (Daclatasvir and Letermovir) and one antibiotic (Rifampicin) dock strongly with both the SARS-CoV-2 nucleocapsid RNA binding domain, but also with the RNA binding site of the DENV capsid protein [39], which is about 80% conserved across all four DENV serotypes. This finding suggests that both the DENV and SARS-CoV-2 nucleocapsid share some common structural features. This is particularly relevant, given that, according to a recent study by Edridge et al., in addition to the coronavirus’ spike protein which elicits neutralizing antibodies, this virus family’s nucleocapsid is recognized as being immunogenic and a “sensitive protein to monitor seasonal coronavirus infections” [40]. Moreover, the nucleocapsid structure is conserved across coronaviruses, including SARS-CoV-2 [40]. Altogether, these observations raise the possibility that an immunological cross-reaction between DENV serotypes and SARS-CoV-2 could be mediated, at least in part, by antibodies (but also a cellular reaction) produced against these two viruses’ nucleocapsid. According to our hypothesis, such an immunological response to DENV could offer some level of protection against the SARS-CoV-2 infection. Obviously, such a hypothesis needs to be tested in a series of further studies.

Interestingly enough, a recent review by Henrina et al. [41] also highlights the enormous similarities between COVID-19 and dengue fever, not only in terms of clinical presentation, but more strikingly, in terms of their common pathophysiology (see Table 2 of Henrina et al., 2020). For instance, the so called cytokine storm syndrome, as well as widespread endothelium dysfunction, is present in severe cases of both COVID-19 and in the most serious clinical manifestation of dengue, known as dengue hemorrhagic fever [41]. Moreover, in both diseases, albeit through different mechanisms, D-dimer levels are elevated [41], and may serve as an indicator of the severity of the clinical manifestation [41].

**Table 1.**
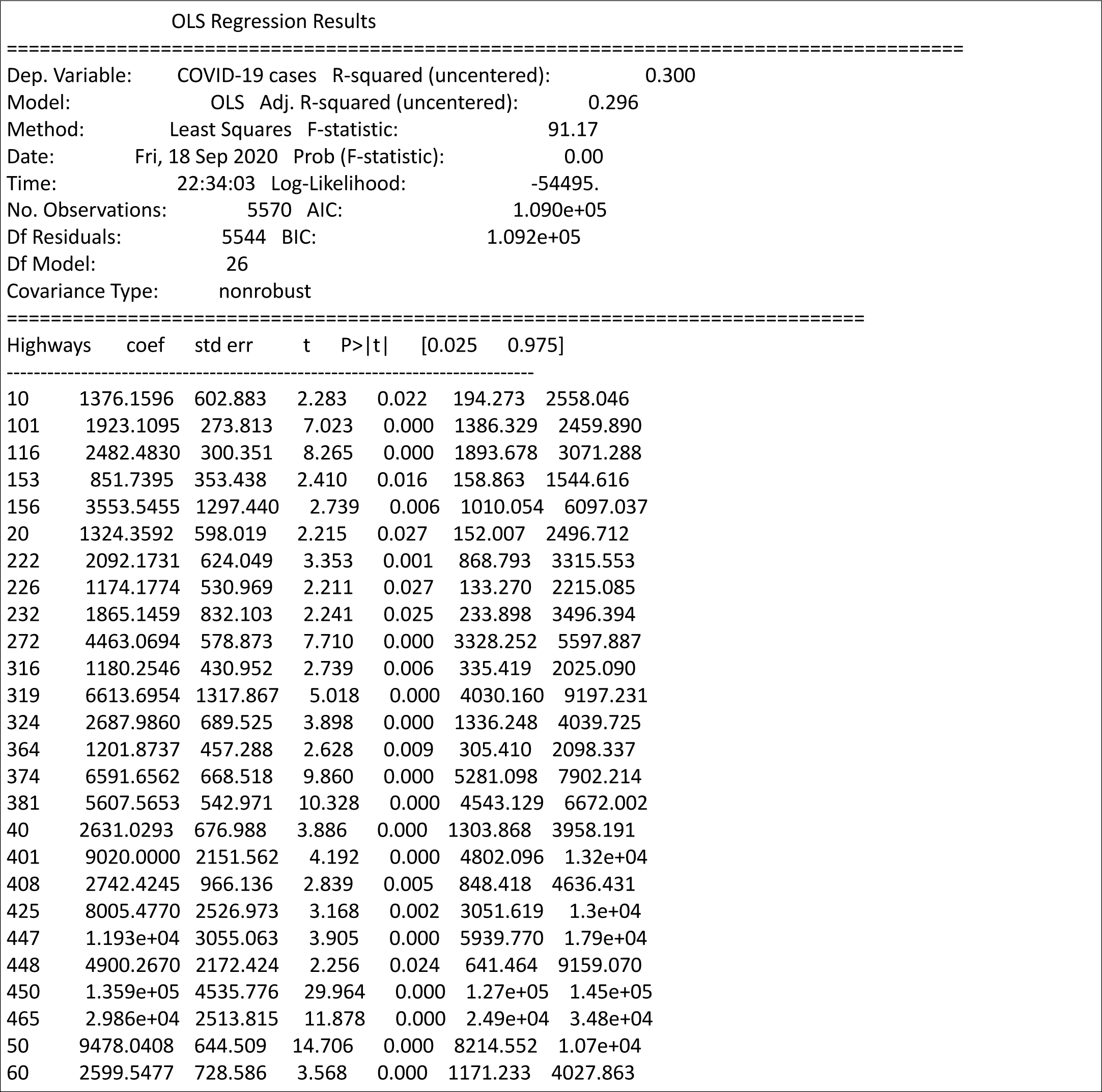
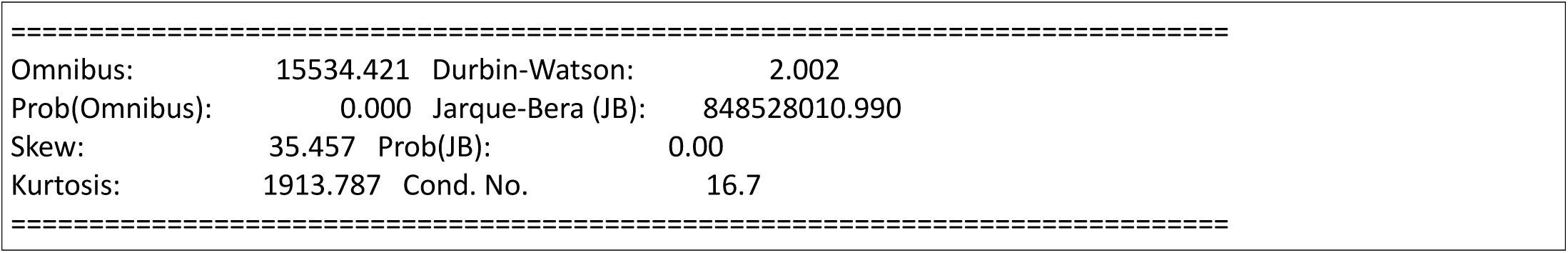
Output of multivariate linear model using federal highways as independent variables, defined at city level with 1 where it crosses and 0 where not, and COVID-19 accumulated cases up to the 12^th^ of September as dependent variable. Software used: Python/stamodels.

**Table 2.**
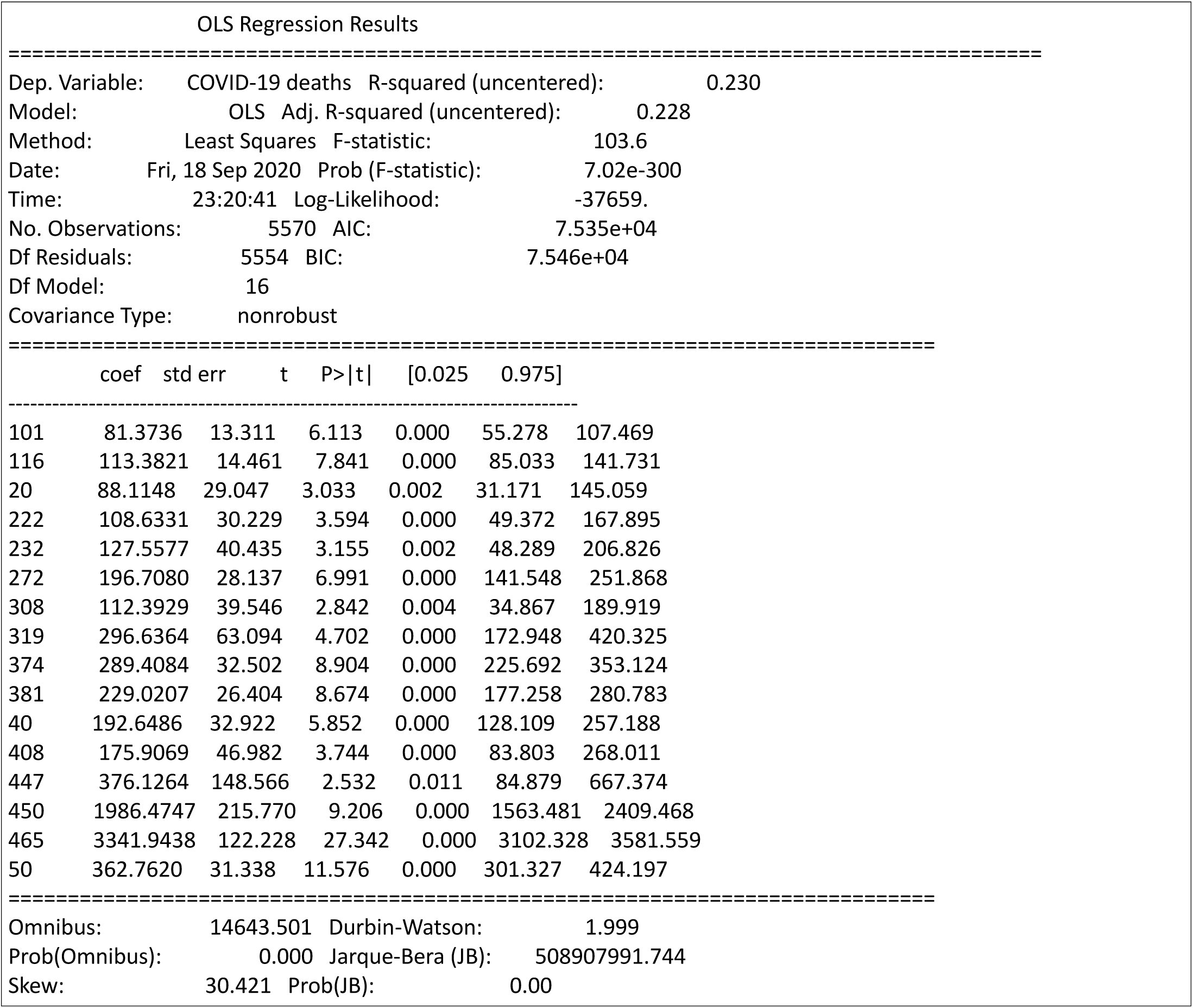
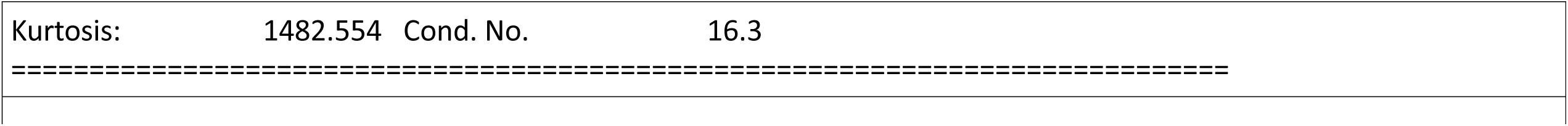
Output of multivariate linear model using federal highways as independent variables, defined at city level with 1 where it crosses and 0 where not, and COVID-19 accumulated deaths up to the 12^th^ of September as dependent variable. Software used: Python/stamodels.

**Table 3.**
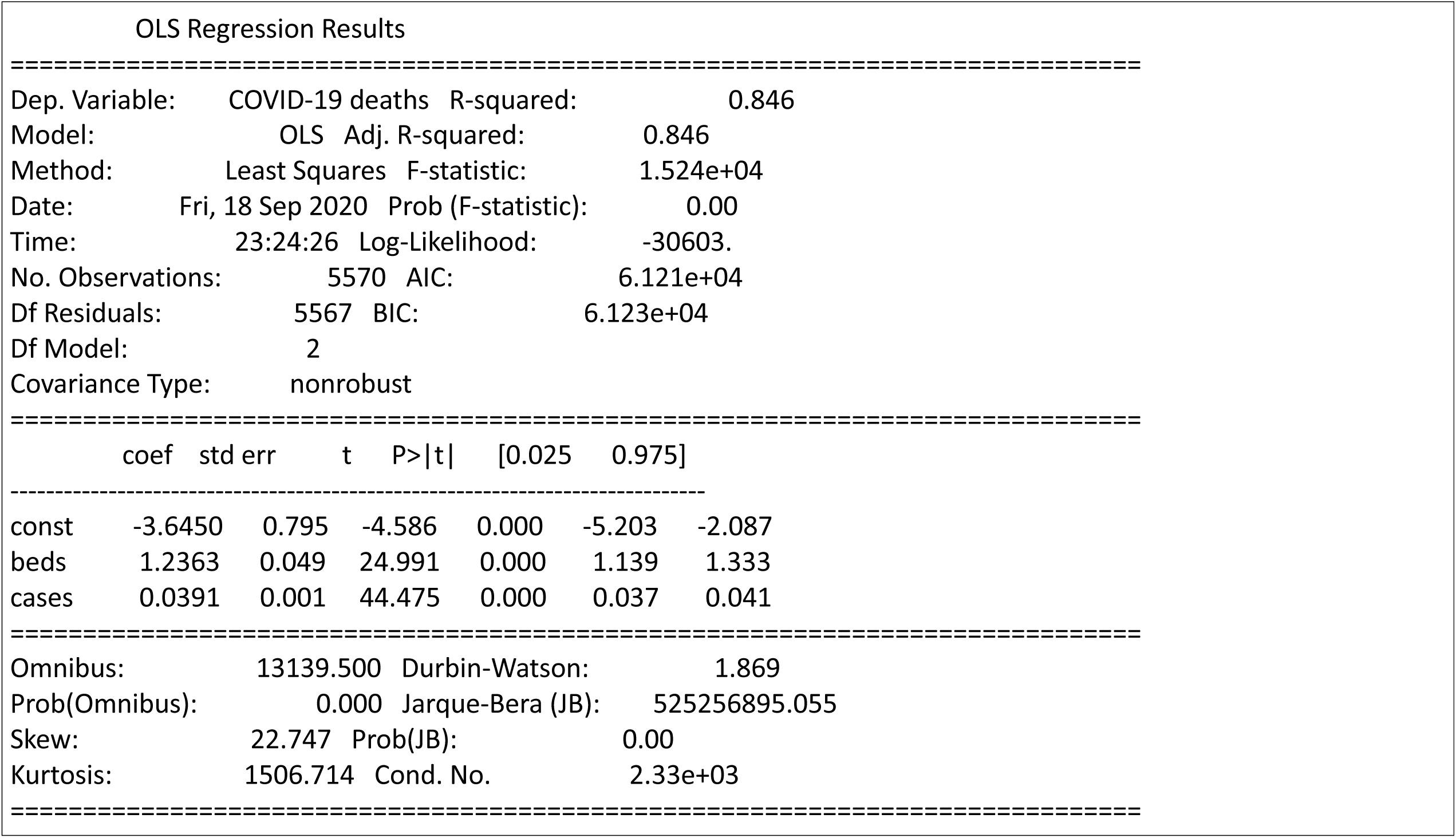
Output of multivariate linear model using hospital beds and accumulated COVID-19 cases (until 1^st^ of July, 2020) as independent variables, and COVID-19 accumulated deaths as dependent variable. Software used: Python/stamodels.

In the context of the worldwide health emergency created by the current coronavirus pandemic, our surprising results bring up the possibility that antibodies (or lymphocyte B and T cells) produced as a response to an episode of dengue fever may provide a certain level of immunological protection against COVID-19. That immediately raises the intriguing hypothesis that immunization for dengue fever could also induce some level of clinically significant immunological protection against SARS-CoV-2. Although many further studies will certainly be needed to clarify whether this is a valid theory and how high and long lasting such a protection could be, there are several factors that speak in favor of pursuing this line of inquiry as a follow-up to the findings reported here. First, currently there are at least two potential dengue vaccines either approved or in final stages of Phase III clinical trials [42, 43]. Dengvaxia (Sanofi, Paris, France), also known as CYD-TDV, was approved by the World Health Organization in 2016, and later licensed in more than 20 countries, after positive results on Phase III clinical trials that showed levels of up to 66% efficacy overall, but only 38% for seronegatives, among children tested in Asia and Latin America [43]. However, a longer follow up of Dengvaxia’s effects revealed that children who were seronegative for DENV exhibited a higher than expected chance of developing serious dengue symptoms that required hospitalization [44]. That triggered a revision of the vaccine’s recommendation, limiting its use to people who had already been exposed to dengue and showed antibodies to at least one dengue serotype. Despite this limitation, the European Medicines Agency and the US Federal Drug Administration (FDA) approved Dengvaxia for use in December 2018 and in May 2019 respectively, provided it is preceded by immunological profiling and given only to seropositive subjects.

A second vaccine, TAK-003 (TAKEDA) has already reported partial results on its Phase III clinical trials [42]. Out of 13,380 subjects that received at least one dose of TAK-003, the vaccine’s overall efficiency reached 80.9%, and 75% in seronegatives [42]. When dengue hospitalizations were considered, TAK-003’s efficiency reached 95.4% of the tested population [42]. TAK-003 was more effective (97.7%) for the DENV-2 serotype, but also moderately effective for the other three (73.7% for DENV-1, 63.2% for DENV-3, and inconclusive results of 63% for DENV-4). Finally, different from DengvaxiaTAK-003, Phase III data revealed an incidence of severe adverse effects similar to the control group [42].

Given this context, we propose that the next step would be to carry out clinical studies to measure how effective infection by DENV has been in protecting patients against SARS-CoV-2 infection in a population in which DENV was very prevalent during 2019-2020. Such a clinical study could quickly yield some fundamental information and test our hypothesis in a relative short-time. Assuming that our hypothesis is confirmed, i.e. that DENV infection produces a clinically relevant level of COVID-19 immunity, consideration could be given to the next step: testing whether immunization with a dengue vaccine can lead to similar levels of protection against COVID-19. If these second level studies confirm our hypothesis, one could imagine using a safe and efficacious dengue vaccine, on an emergency basis, to reduce the transmission rate of SARS-CoV-2, by producing a significant level of immunity before a specific vaccine to SARS-CoV-2 becomes available.

The multiple-step course of action proposed here would be totally justified on both scientific and ethical grounds provided a few key prerequisites are fulfilled. Firstly, dengue fever is a serious disease that can lead to hospitalization and even death, particularly in those infected a second time [24]. Therefore, immunizing large cohorts would be totally justified, provided that a safe and efficacious vaccine is available. That means finding a definite solution for avoiding serious side effects in the population that is seronegative for DENV in countries where dengue fever is endemic, like Brazil, and most of Latin America, and Asia. Secondly, in countries where dengue is not present, dengue immunization does not offer any major risk since subjects are highly unlikely to be brought in contact with dengue. As such, dengue immunization could proceed, as soon as definitive clinical evidence shows categorically that our hypothesis is meritorious. Optimism about this proposal is reinforced by the fact that the DENV and SARS-CoV-2 belong to distinct virus families. Therefore, the chances that those immunized against dengue may later develop serious side effects when exposed to SARS-CoV-2, such as those mediated by the phenomenon of antibody dependent enhancement [45], should be, at least in theory, much smaller, or even non-existent, than those observed when closely-related viruses are involved (like the DENV and Zika mediating viruses, both members of the Flavivirus family) [46].

Finally, it is worth speculating that, since other flaviviruses exist that generate human diseases, like Zika and yellow fever for instance, still unknown cross-immunological interactions between SARS-CoV-2 and other flaviviruses (or even other viruses, for that matter) may exist. Investigating and deciphering such interactions may be highly relevant for establishing other potentially useful emergency mitigation strategies for reducing the rate of growth of SARS-CoV-2 infection. For instance, since the traditional yellow fever vaccine is well-known and readily available worldwide, it would be interesting to investigate whether there is any cross-immunological interaction between the yellow fever virus and SARS-Cov-2 in subjects recently vaccinated against yellow fever. In case this is true, the yellow fever vaccine could also be considered as an emergency strategy to reduce the number of cases of COVID-19.

As emphasized above, this and other potential emergency strategies will require further clinical studies to decide whether they constitute valid and safe clinical approaches to mitigate the human impact of the COVID-19 pandemic while there is no approved vaccine or therapy for dealing with SARS-CoV-2.

## MATERIALS AND METHODS

### Data sources for COVID-19 cases and deaths

We obtained data describing the temporal evolution of COVID-19 cases and deaths in Brazil at the municipal and state levels from several sources, including the Brazilian Ministry of Health [47], official daily epidemiological bulletins issued by each Brazilian State [48], and other souces as compiled by Cota et al. [49]. Both cases and death data refer to notifications per day. To compute incidence (cases per 100,000 inhabitants), we used population size estimates for each of the 5,570 Brazilian municipalities for 2019 [50]. Such population size estimates were aggregated to allow the computation of COVID-19 at the state level.

We also used the Ministry’s data on Severe Acute Respiratory Infections (SARI) data, in which COVID-19 cases represents close to 98% of the data in 2020, to obtain detailed information about patients’ residence and hospitalization location throughout Brazil (https://opendatasus.saude.gov.br/dataset/bd-srag-2020). The SARI data contains only a subset of the official reported COVID-19 cases since they cover only hospitalized cases. Dengue epidemiological and serological data were provided by data published in the official epidemiological bulletins regularly during 2019 and 2020 by the Brazilian Ministry of Health.

### Data source for the Brazilian Federal road system

The shapefile with geospatial data describing the distribution of the Brazilian federal roads was obtained from the Brazilian National Road System [51]. Roads were categorized according to the official typology: longitudinal roads (codes starting with 1, as in BR101) are those crossing the country from north to south; transversal roads which cross the country from east to west (codes starting with 2); diagonal roads (codes starting with 3); connector roads are shorter roads connecting major federal roads (codes starting with 4); and radial roads, those departing from Brazil’s capital, Brasilia, which has a central geographical position (codes starting with 0).

### Highway multi-linear model

To investigate the most representative highways concerning the COVID-19 spatial distribution pattern, we built a multi-linear model on a city level. Highways were included in the model as dummy variables, considering 1 for cities it crosses, and 0 for cities it does not cross. Model selection started from all federal highways and then a 3 step filtering process was performed. The first filter eliminated variables (representing highways) with coefficients with statistical p-values larger than 0.10, then 2 subsequent steps of elimination were performed for variables with p-values larger than 0.05. The resulting multi-linear model significantly adjusted 26 highways (R2 = 0.3, p < 0.025 for all variables) when the response variable was the accumulated COVID-19 cases on the 12th of September 2020, and significantly adjusted 16 highways (R2 = 0.23, p < 0.015 for all variables) for accumulated COVID-19 deaths on the same date.

### Spatial spreading model

The spatial spreading of COVID-19 throughout the country was modelled following the approach described in Peixoto et al. [7]. This approach is based on a complex mobility network of all Brazilian cities coupled with a compartmental model containing infected and susceptible individuals, adequate for simulations of initial epidemic dynamics. The mobility data is based on individual pairwise mobile geolocation data, resulting in multiple daily travel information between cities, collected from the Brazilian company Inloco (https://www.inloco.com.br/covid-19). The compartmental model adopted an infection rate of r=0.2 individuals per day, because it provides more realistic forecasts for the initial growth of the pandemic in Brazil, compared to the initial infection rates obtained for the country (e.g. [22, 52].

The flux intensity parameter was set to s=1, that is, no compensation of the flux intensity was performed, and the real daily sampled movement counts were used to infer the mobility between cities. The code and mobility data are available at the GitHub repository https://github.com/pedrospeixoto/mdyn

### Model of the super-spreaders

For each state capital of Brazil, a separate simulation considering one infected individual in the capital was performed using the spatial spreading model. The simulations were run from 2020-03-01 until 2020-05-01, with a result, on the final day, consisting of the potential spatial spreading pattern for each capital city. The super-spreaders model was built by projecting, in the least-squares sense, the daily observed COVID-19 cases into the sub-space generated by the linear combination of the spreading patterns obtained for each capital city. This provided a linear model for each day of observed COVID-19 cases, with coefficients representing the degree of participation of a given city in the observed spatial pattern of COVID-19 cases. With a basis of the most representative 17 capital spreading patterns, the super-spreaders linear model accounted for at least an adjusted coefficient of determination of 0.94 in all dates analyzed in this study (over 0.98 in the first 2 months analysed).

### Model prediction of case spatial distribution

The spatial spreading model was used to infer the theoretical spatial distribution of initial cases across all states in Brazil. The model was initialized with the observed COVID-19 cases for each municipality in the country on March 30^th^ of 2020, the day when all international Brazilian airports were closed. From this day on, only regional dissemination of COVID-19 cases followed. While the model is not expected to capture the exact number of cases in future times, it provides an accurate estimate for the geographic spread and distribution of cases, which was by then mostly dominated by the mobility data.

### Flavivirus correlation analysis

To ensure robustness, the interplay between COVID-19 and Flavivirus epidemics was analyzed considering several different viewpoints. Simple linear regression analysis was performed in most cases, considering exponential growth where appropriate. Also, for each state of the country, a log-log linear model was adjusted considering the period that goes from the observation of the 10th COVID-19 cases until 90 days afterwards. These models were adherent with at least a coefficient of determination of 0.95, providing a very good representation of the initial growth of the pandemic in each state. The slopes of these models, representing the initial growth rate of the COVID-19 epidemic in Brazil, were used in the correlation analysis with the population levels of IgM for dengue fever in each state.

### Methodological limitations

The dynamical model, while fully coupled in space by mobility, adopts a simplified compartmental dynamic, with susceptible-infected only. This limits the model’s ability to foresee longer periods in time, compared for instance with the many existing variants of SEIR models. However, it reduces the complexity in parameter calculations and, most importantly, in the estimation of initial conditions for unobserved compartments. To compensate for this limitation, we only used the model for short periods of time and focused our conclusions on the spatial distribution patterns of the forecasted results rather than on the precise case count calculated.

### Dataset information and limitations

Mobility data: The mobile mobility dataset was provided by Inloco (https://www.inloco.com.br/covid-19), available upon request, and samples approximately one-fifth of the Brazilian population. While having a vast coverage of the Brazilian population, it may have uneven distribution in space, age, and social classes. The data is mainly dominated by adults and, is more abundant in large cities, but samples more than 90% Brazilian municipalities. This dataset is, to the best of our knowledge, the largest database of the kind available for research in Brazil.

## Data Availability

All datasets used in this study are publicly available and the references or links to the data sources are provided in the text.

## Acknowledgments

MALN was supported by a Duke University Medical Center Distinguished Professor Endowed Chair. CSA and RLGR were supported by the Brazilian Synthesis Center on Biodiversity and Ecosystem Services (SinBiose/CNPQ). PP was supported by a research grant from Fundação de Amparo à Pesquisa do Estado de São Paulo (FAPESP 16/18445-7).

## Author Contributions

MALN designed the study, collected and analyzed data, identified the “boomerang effect discovered the interaction between COVID-19 and dengue fever, and wrote the paper. RR helped design the study, executed all the highway and “boomerang effect” studies, participated in all data analysis, and helped in the manuscript writing. PP was responsible for all mathematical models and statistical analysis of the manuscript and participated in the manuscript writing. CA was responsible for data collection and analysis of the hospitalization data throughout Brazil and in the characterization of the boomerang effect. She also contributed to the writing of the manuscript. All authors discussed approved all the arguments provided in the paper.

